# Genetic Insights into the Relationship Between Psychiatric Disorders and Irritable Bowel Syndrome: A Mendelian Randomization Analysis

**DOI:** 10.1101/2024.02.09.24302578

**Authors:** Mahmud Omar, Mohammad Omar, Reem Agbareia, Saleh Nassar

## Abstract

**Background:** Irritable Bowel Syndrome (IBS) is often accompanied by psychiatric conditions, yet the causal relationship remains uncertain. This study leverages Mendelian Randomization to explore the genetic basis of the association between IBS and various psychiatric disorders.

**Methods:** We analyzed GWAS data to assess the causal effects of Major Depressive Disorder (MDD), Anxiety, and other psychiatric disorders on Irritable Bowel Syndrome (IBS). Stringent criteria were used to select genetic instrumental variables, and we applied multiple Mendelian Randomization (MR) methods, including a reverse MR analysis, to investigate the impacts comprehensively.

**Results:** Our study found a significant causal relationship between IBS and MDD (IVW OR: 1.328, 95% CI: 1.122, 1.572, P=0.001) and a slight but significant link with Anxiety Disorders (IVW OR: 1.0611, 95% CI: 1.0184, 1.1056, P=0.0046). Bipolar Disorder, Schizophrenia, OCD, ADHD, Anorexia Nervosa, and Insomnia did not show significant causal connections with IBS. The reverse MR analysis demonstrated a positive correlation between IBS and MDD (IVW OR: 1.522) and a negative one with ADHD (IVW OR: 0.866), while findings for Schizophrenia and other disorders were not significant.

**Conclusion:** The study confirms a unidirectional causal relationship between IBS and certain psychiatric disorders, highlighting the relevance of the gut-brain axis. These insights contribute to the understanding of IBS pathophysiology and underscore the need for considering mental health in IBS management.

## Introduction

Irritable Bowel Syndrome (IBS) is a common gastrointestinal disorder characterized by symptoms such as abdominal pain, bloating, and altered bowel habits (1). Its etiology remains complex and multifactorial, with interactions between gut-brain axis, environmental, and genetic factors (2). Notably, IBS significantly impacts quality of life and imposes a substantial burden on healthcare systems (3).

The association between IBS and psychiatric morbidity has been a focal point in recent research. Studies have consistently observed higher rates of psychiatric disorders, such as anxiety and depression, among individuals with IBS compared to those without (4–8). However, the nature of this relationship remains unclear (9–11). While some suggest that psychiatric conditions may exacerbate IBS symptoms, others propose that IBS might predispose individuals to psychiatric morbidity (4,5,8,10,11).

Our study aims to clarify the causal dynamics between various psychiatric disorders and IBS using Mendelian Randomization (MR). This will be accomplished through an extensive examination of the genetic causal link between IBS and various psychiatric disorders: Major Depression (MDD), Bipolar Disorder, Anxiety, Schizophrenia, Obsessive Compulsive Disorder (OCD), Attention Deficit Hyperactivity Disorder (ADHD), Anorexia Nervosa, and Insomnia.

This method leverages genetic variants as proxies to estimate the causal effect of an exposure (in this case, psychiatric disorders) on an outcome (IBS), while minimizing confounding factors and reverse causation (12,13). By analyzing data from large-scale Genome-Wide Association Studies (GWAS), we seek to provide new insights into whether psychiatric disorders are a contributing cause of IBS or merely co-occurring conditions. This understanding could have significant implications for both the treatment and management of IBS, and the broader understanding of its pathophysiology.

## Materials and Methods

### Mendelian Randomization Study

Our study leverages Mendelian Randomization (MR) to assess the potential causal relationship between various psychiatric disorders and IBS (Figure 1). We ensured adherence to the standard MR assumptions: genetic instrumental variables (IVs) must be strongly associated with the psychiatric disorders (association), must not be associated with confounders (independence), and should affect IBS only through the psychiatric disorders (exclusivity). We also evaluated the possibility of pleiotropy, where genetic variants might influence the outcome through pathways other than the exposure.

**Figure 1:**
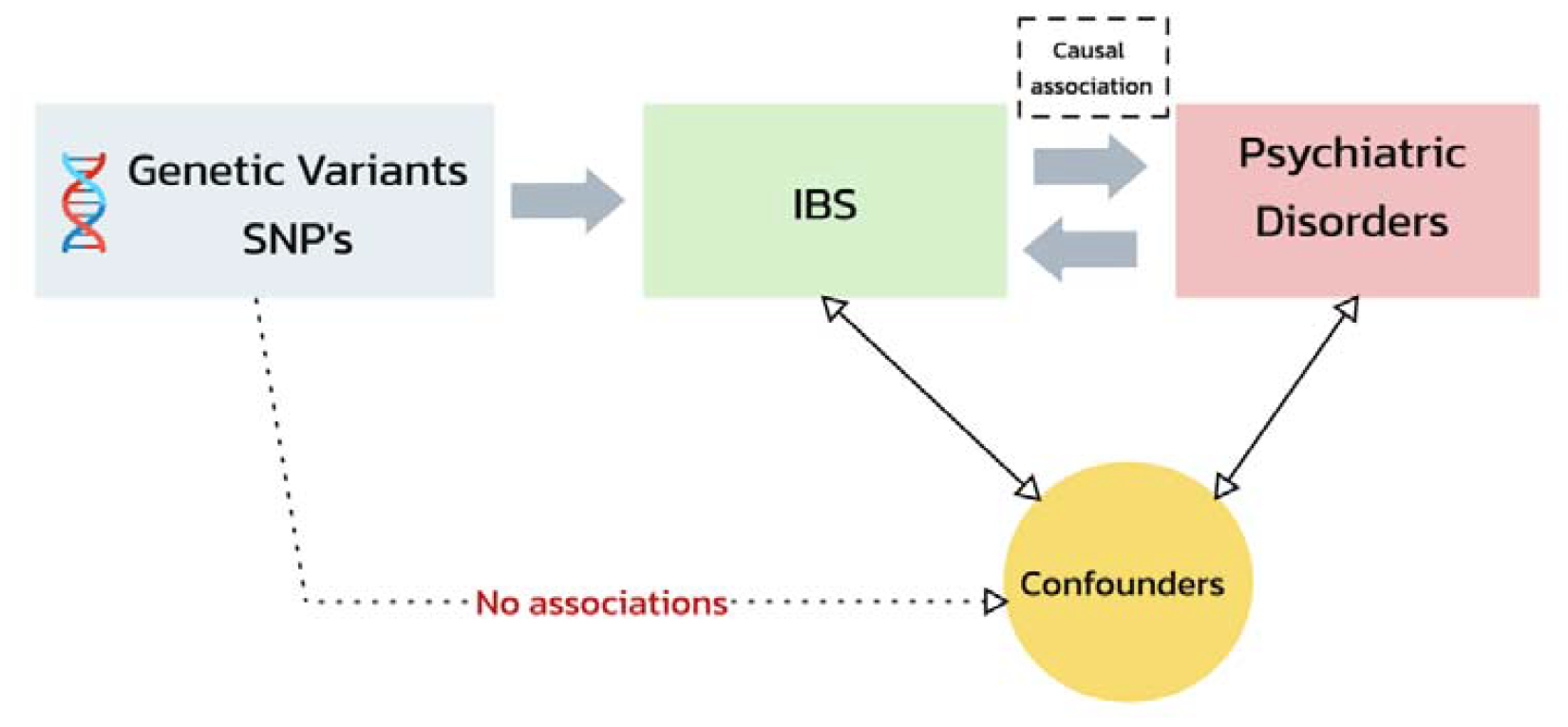
Flowchart of Mendelian Randomization Analysis: IBS and Psychiatric Disorder Associations.

### Selection of Instrumental Variables

Instrumental variables were carefully chosen from Genome-Wide Association Studies (GWAS) datasets. For IBS, we used data from a GWAS consisting of 53,400 cases and 433,201 controls, summing up to a total sample size of 486,601 with 9,739,966 SNPs (14). The outcomes analyzed included MDD, Bipolar Disorder, Anxiety, Schizophrenia, OCD, ADHD, Anorexia Nervosa, and Insomnia, with participants primarily of European descent. Summary-level data for these outcomes were obtained from the IEU GWAS database. Table 1 shows the detailed information on the outcome datasets. Selection criteria for SNPs included a genome-wide significance threshold of p < 5 × 10^-8 and the exclusion of those in linkage disequilibrium (LD r^2 = 0.001 within 10,000KB). Minor allele frequencies (MAF) were considered only if above 0.01. When necessary, proxy SNPs were utilized, and palindromic SNPs with MAF > 0.42 were excluded. The instrumental strength of SNPs was quantified using an F-statistic, with an F-value of 10 as the minimum threshold to ensure the validity of the IVs. We harmonized all instrumental variables for each trait to ensure the genetic associations reflect the same effect allele. Statistical Methods

**Table 1:**
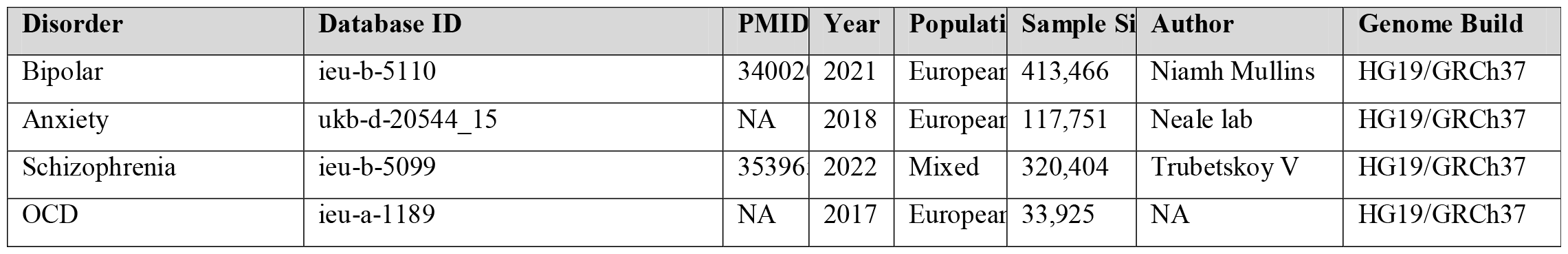
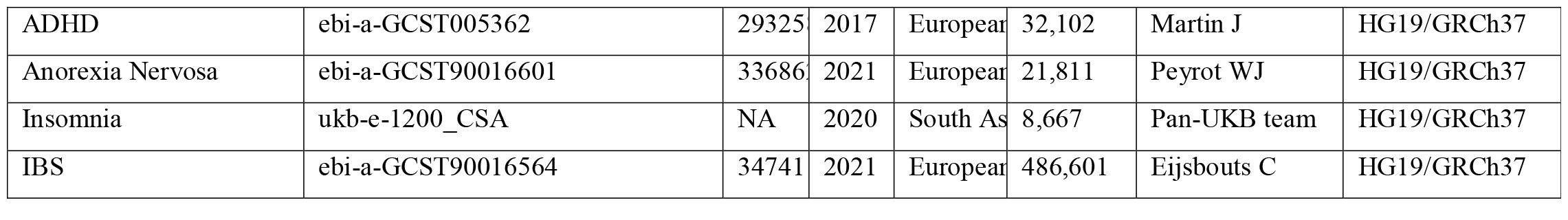
Summary data.

The analysis utilized the inverse variance weighted (IVW) method as the primary estimator of the causal effect, under the assumption that all SNPs are valid IVs. To handle the potential of pleiotropy, we opted for the random-effects IVW method over fixed-effects. Additional methods including MR-Egger regression, Weighted Median Estimator (WME), Weighted Mode, and Simple Mode were implemented to provide a comprehensive assessment of the MR estimates’ robustness. Cochran’s Q-tests were conducted to examine heterogeneity among the IVs, and MR-Egger regression was used to test for horizontal pleiotropy. We set a p-value cutoff of 0.05 to determine the significance of the causal estimates. The statistical analyses were conducted using the TwoSampleMR and MRPRESSO packages within the R software environment (Version 2023.03.0+386) (15).

### Reverse Mendelian Randomization Analysis

The reverse MR analysis explored psychiatric disorders as exposures influencing the risk of developing IBS. This analysis followed the same methodological framework as the primary MR analysis without the need for a lenient significance threshold or relaxed LD criteria for any of the psychiatric conditions studied. Valid IVs for each psychiatric disorder were identified and used to estimate their effect on IBS.

## Results

### Major Depressive Disorder (MDD)

For MDD, our MR analysis revealed a modest association with IBS. The Inverse Variance Weighted (IVW) method yielded an odds ratio (OR) of 1.328 (95% CI: 1.122, 1.572, P=0.001), indicating a statistically significant effect. The Weighted Median estimate corroborated these findings with an OR of 1.279 (95% CI: 1.093, 1.498, P=0.002). Sensitivity analyses, including MR-Egger regression and MR-PRESSO, did not indicate the presence of horizontal pleiotropy, thus reinforcing the validity of our findings (MR-Egger intercept P=0.1561, MR-PRESSO Global Test P=0.174) (Figures 2-4).

**Figure 2:**
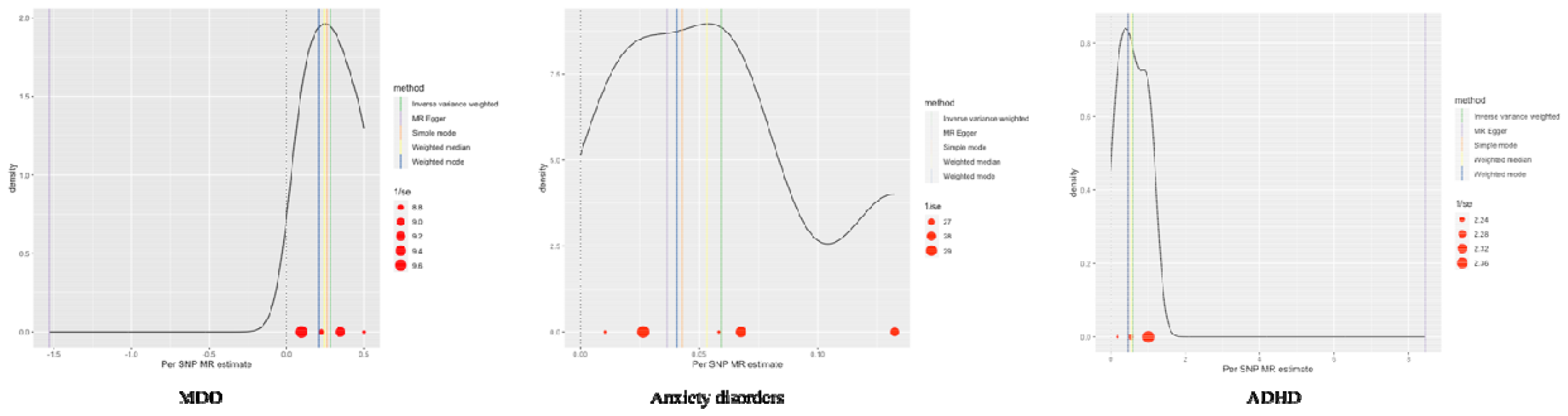
Density Plots of SNP-Based MR Estimates for MDD, ADHD, and Anxiety Disorders.

**Figure 3:**
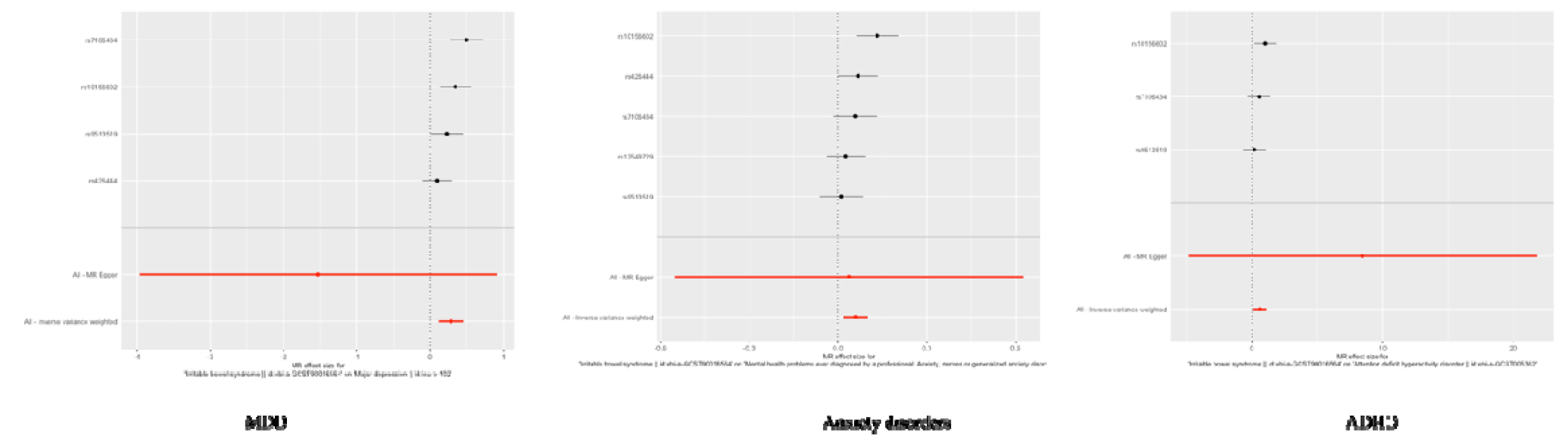
Mendelian Randomization Analysis: Causal Effects of MDD, ADHD, and Anxiety Disorders on IBS.

**Figure 4:**
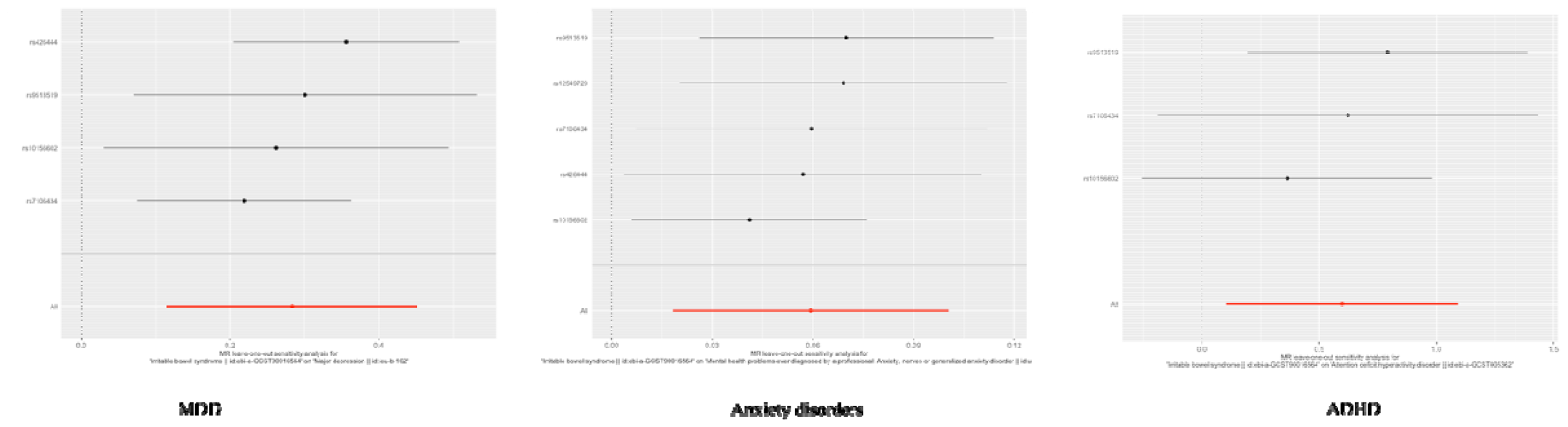
Leave-One-Out Sensitivity Analysis for MR Estimations on IBS with MDD, ADHD, and Anxiety Disorders.

### Bipolar Disorder

The causal effect of Bipolar Disorder on IBS was not statistically significant in the primary analysis, with the IVW method producing an OR of 1.2265 (95% CI: 0.6789, 2.2159, P=0.4987). Sensitivity tests supported the null finding, showing no evidence of pleiotropy or outliers that could bias the results (Pleiotropy Test P=0.2380, MR-PRESSO Outlier Test identified outliers).

### Anxiety Disorders

For Anxiety Disorders, the MR analysis showed a slight significant association with IBS. The IVW estimate was OR=1.0611 (95% CI: 1.0184, 1.1056, P=0.0046), which remained significant across other MR methods, including the Weighted Median (OR=1.0547, 95% CI: 1.0097, 1.1017, P=0.0167). Sensitivity analyses did not alter the inference, suggesting robustness in the association (MR-PRESSO Global Test P=0.389) (Figures 2-4).

### Schizophrenia (SCZ)

The MR estimates for Schizophrenia provided suggestive, yet non-significant results with the IVW method yielding an OR of 1.117 (95% CI: 0.775, 1.610, P=0.5542). The Weighted Median method indicated a non-significant effect (OR=0.890, 95% CI: 0.691, 1.145, P=0.3633). Heterogeneity and pleiotropy analyses did not reveal any significant biases.

### Obsessive-Compulsive Disorder (OCD), ADHD, Anorexia Nervosa, and Insomnia

For OCD, ADHD, Anorexia Nervosa, and Insomnia, our MR analyses provided non-significant causal estimates with large confidence intervals, suggesting that the evidence does not support a strong causal effect of these conditions on IBS. Notably, the analysis for ADHD showed a significantly inflated OR due to the MR Egger method (OR=4665.97, CI: 0.0075, 2888182798.92, P=0.4316), which likely results from horizontal pleiotropy or other biases. Table 2 summarizes the MR results across the different methods, and Table 3 summarizes the results of the sensitivity and complementary tests for each disease.

**Table 2:**
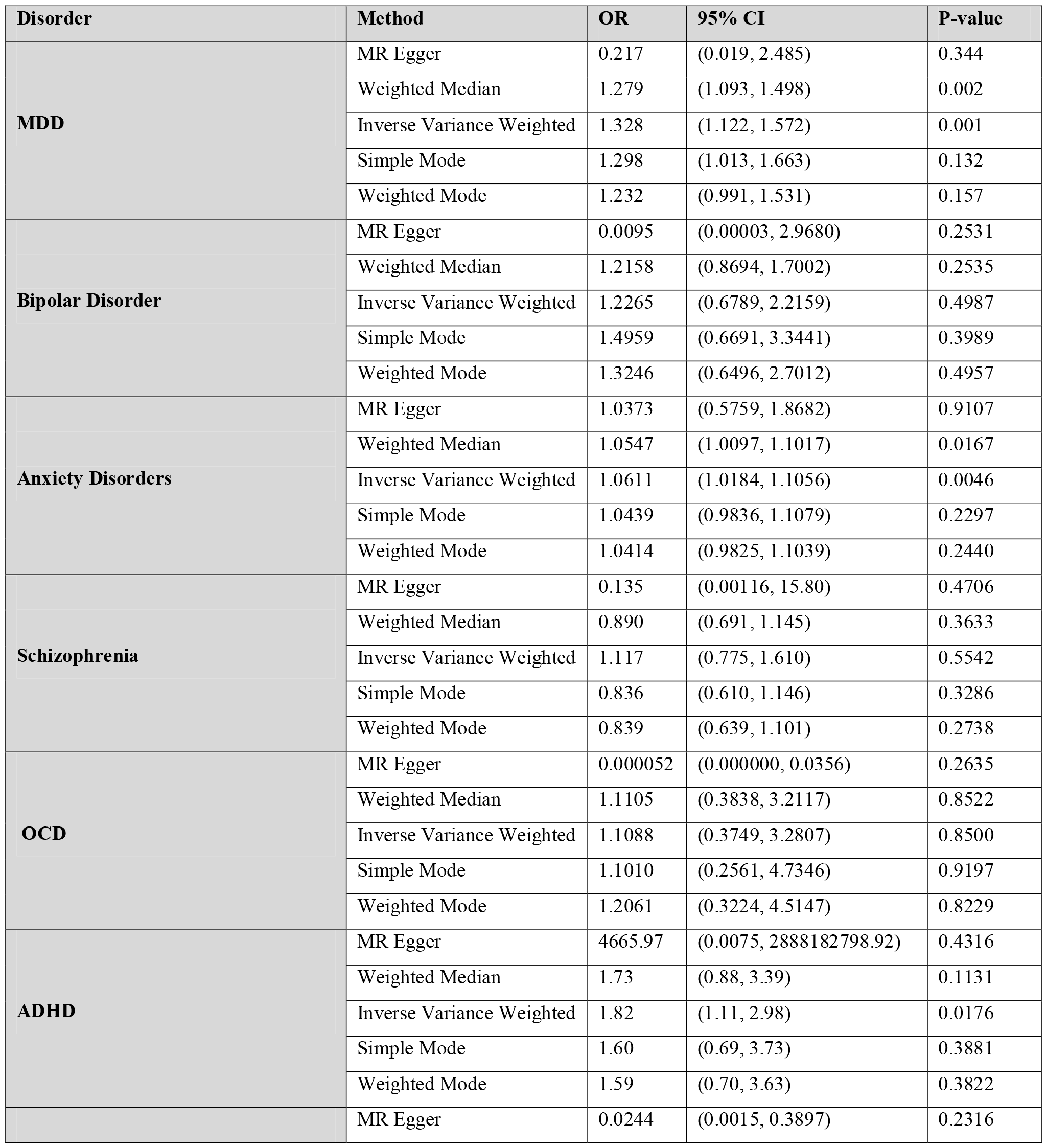
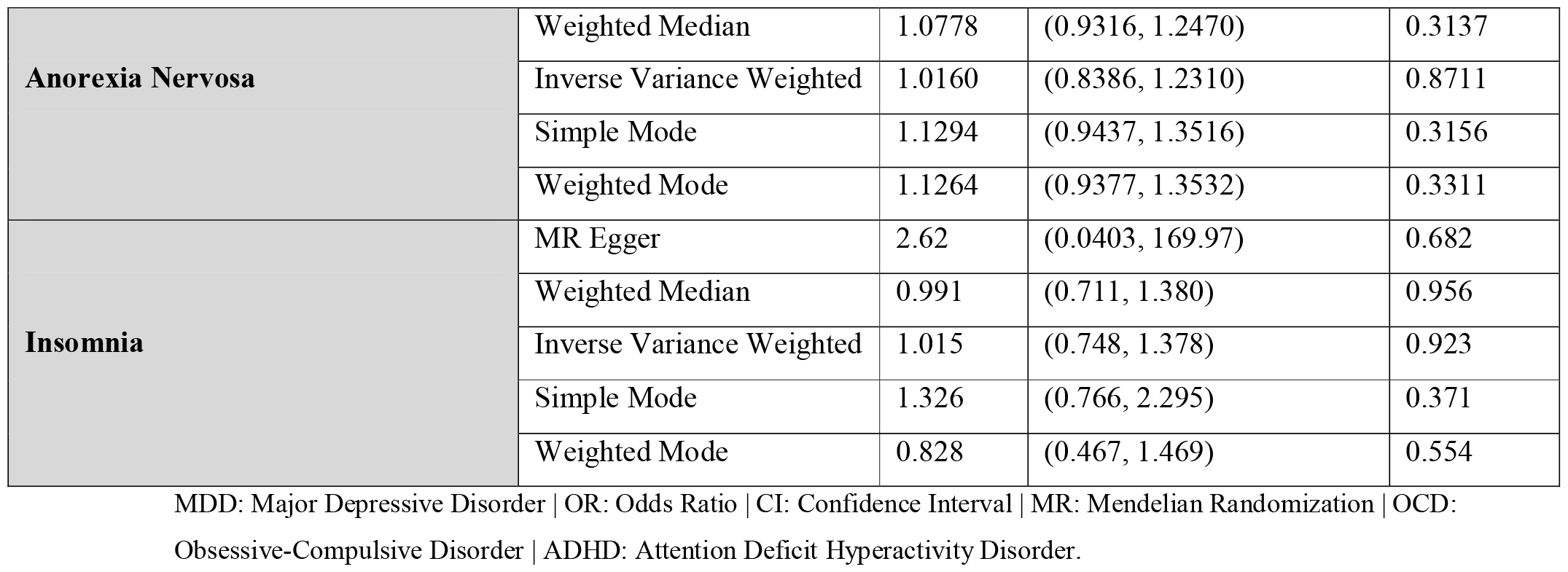
Causal Estimates for Psychiatric Disorders and IBS.

**Table 3:** Summary of Mendelian Randomization Analyses Across Psychiatric Disorders.

| Disorder | Heterogeneity Test (Cochran's Q; Egger; IVW) | P Value | Leave One Out | MR-PRESSO (Global Test; Outliers) | MR Egger Regression (Intercept) |
| --- | --- | --- | --- | --- | --- |

|  |  |  |  |  |  |
| --- | --- | --- | --- | --- | --- |
| **MDD** | p=0.1668; p=0.0603 | p | No single SNP significantly altered results | p=0.174; No outliers | p=0.1561 |
| **Bipolar Disorder** | p=0.0197; p=0.0003 | p | No single SNP significantly altered results | p<0.001; Outliers corrected, and statistically different | p=0.2378 |
| **Anxiety Disorder** | p=0.0779; p=0.1452 | p | No single SNP significantly altered results | p=0.389; No outliers | p=0.8810 |
| **Schizophrenia** | p=0.0014; p=0.0006 | p | No single SNP significantly altered results | p<0.001; Outliers corrected, and statistically different | p=0.9057 |
| **OCD** | p=0.4000; p=0.2342 | p | No single SNP significantly altered results | p=0.529; No outliers | p=0.2129 |
| **ADHD** | p=0.4709; p=0.3960 | p | No single SNP significantly altered results | p=0.112; No outliers | p=0.8469 |
| **Anorexia Nervosa** | p=0.9033; p=0.0305 | p | No single SNP significantly altered results | p=0.157; No outliers | p=0.5542 |
| **Insomnia** | p=0.1190; p=0.1818 | p | No single SNP significantly altered results | p=0.283; No outliers | p=0.4572 |

### Reverse Mendelian Randomization Analysis

In the reverse analysis of our Mendelian Randomization study, we examined the relationship between various psychiatric disorders and IBS. For MDD as an exposure, a significant positive association with IBS was observed using the IVW method (OR: 1.522, 95% CI: 1.379 to 1.681, p<0.0001). This finding was supported by other methods including Weighted Median and Weighted Mode. The MR Egger Pleiotropy Test indicated no substantial pleiotropy (Egger intercept: 0.01185023, p=0.145722), and MR-PRESSO results showed significant heterogeneity but no distortion from outliers.

For ADHD, the IVW method revealed a significant negative association with IBS (OR: 0.866, 95% CI: 0.769 to 0.976, p=0.0178). The MR Egger Pleiotropy Test did not suggest significant pleiotropy (Egger intercept: -0.05972416, p=0.3173726), and there was no significant heterogeneity.

Regarding Schizophrenia, our analysis using the IVW method yielded an OR of 1.029 (95% CI: 1.004 to 1.055, p=0.0247). However, this result is within the region of practical equivalence (ROPE), indicating that the association might not be practically significant despite its statistical significance.

Other psychiatric disorders examined in the reverse analysis, including bipolar disorder, anxiety disorders, and anorexia nervosa, did not show significant associations with IBS (Table 4).

**Table 4:**
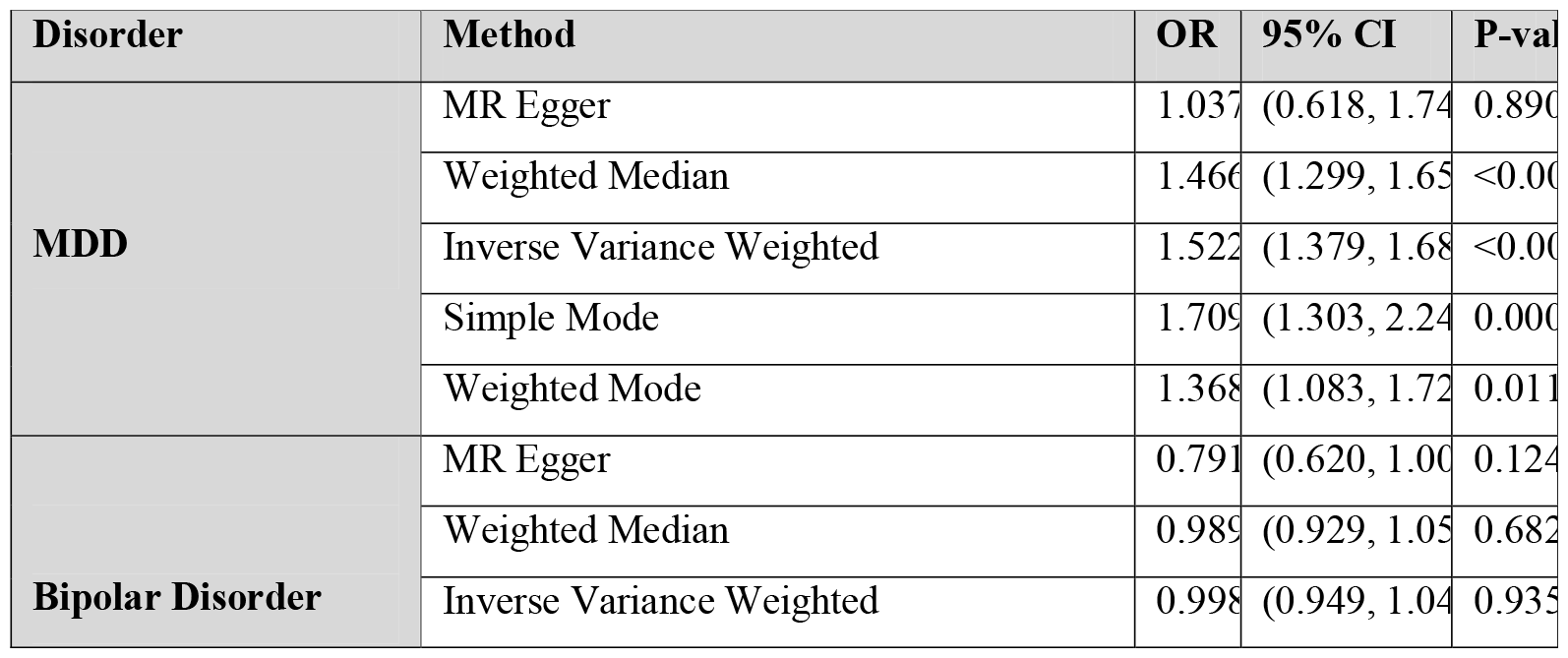
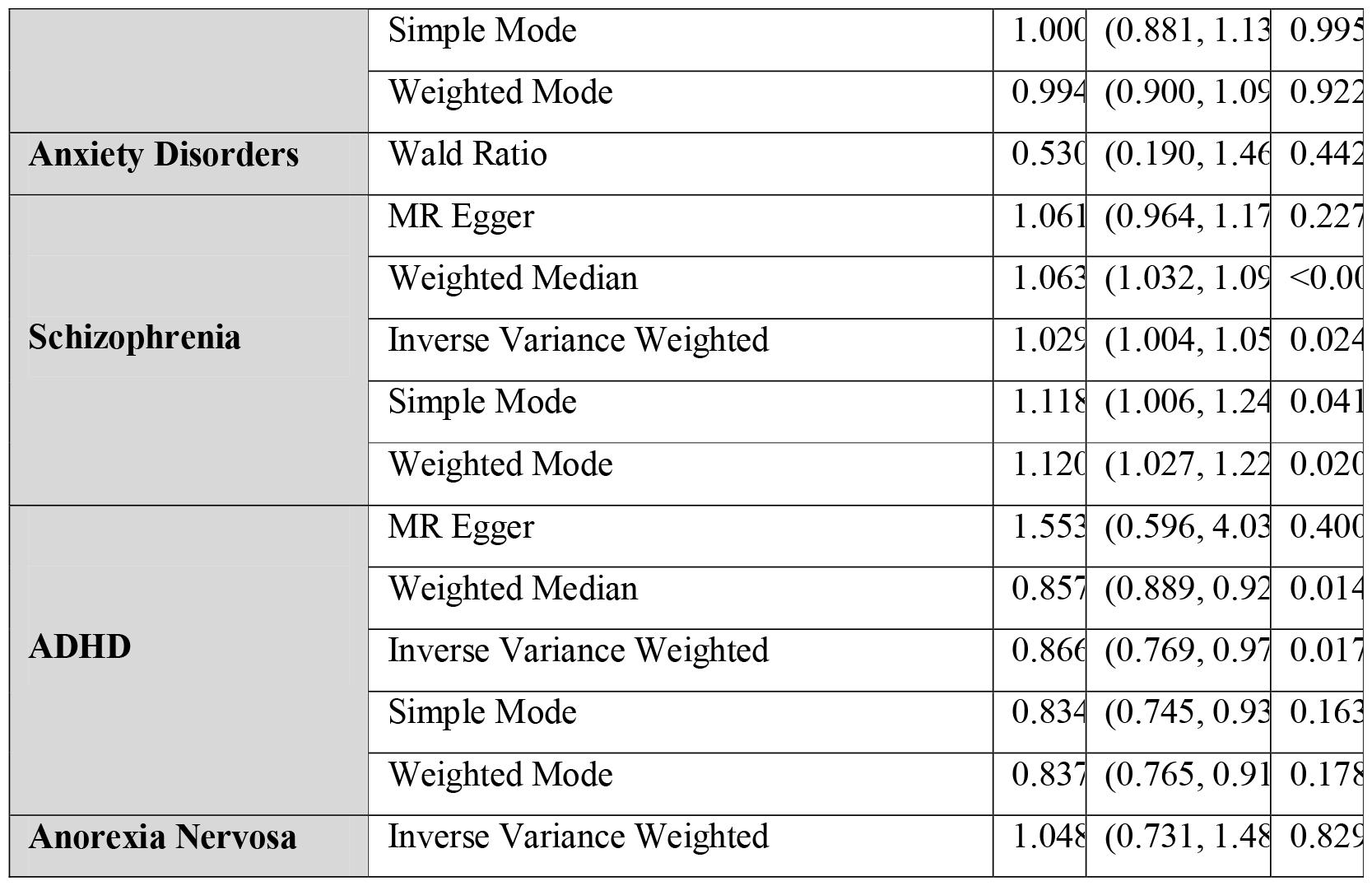
Reverse Analysis: Associations Between Psychiatric Disorders and IBS.

## Discussion

### Summary of Main Results and Comparison to Current Literature

Our study employed Mendelian Randomization to explore the relationship between psychiatric disorders and IBS. We found significant causal associations between IBS and MDD and Anxiety Disorders, while Bipolar Disorder, Schizophrenia, OCD, ADHD, Anorexia Nervosa, and Insomnia showed no significant causal effects. Notably, in the reverse analysis, we found a significant positive association between MDD and IBS, and a significant negative association with ADHD while other psychiatric disorders did not demonstrate significant associations. These results, confirmed by rigorous statistical methods, highlight a specific causal connection between certain psychiatric conditions and IBS, enhancing our understanding of their interaction.

In general, our findings support existing epidemiological and observational studies that identify a clear relationship between IBS and both MDD and Anxiety Disorders (5,16–18). Additionally, the literature generally indicates a lack of a strong connection between IBS and Schizophrenia or Bipolar Disorder, with only a limited number of studies exploring these links (19–21).

### The Role of the Gut-Brain Axis in IBS and Psychiatric Interplay

The modest association of IBS with MDD and Anxiety Disorders, as indicated by our MR analysis, opens new vistas in understanding the gut-brain axis. This finding corroborates many of the current evidence from epidemiological studies that suggest a bidirectional relationship. Specifically, our results align with epidemiological data indicating that IBS increases the risk of developing psychiatric disorders (4), and from a meta-analysis that implies a two-way causal connection (16).

Our Mendelian Randomization (MR) study, compared to Diao et al.’s similar research, highlights key differences possibly stemming from distinct data sources and variables. While Diao et al. identified a causal link between broader psychological distress markers and IBS, our analysis, focusing on specific psychiatric diagnoses like MDD and Anxiety Disorders, found a modest association with IBS (22). This disparity could be attributed to our use of more clinically defined psychiatric conditions versus Diao et al.’s approach, which involved general psychological distress indicators such as visits to a doctor for nerves or anxiety. Moreover, Diao et al. reported no reverse causal relationship from IBS to psychiatric disorders, whereas our study found significant causal effects of psychiatric conditions on IBS (22).

While the established link between IBS and various psychiatric disorders, which significantly impact quality of life and healthcare utilization, is well-documented in existing literature (4,6,17). Our research, suggesting a potential causal connection, further emphasizes the critical role of the gut-brain axis. This axis, involving complex interactions between the central nervous system and the enteric nervous system, plays a key role in the etiology of IBS (17). Factors such as dysregulation in the brain-gut axis, altered gastrointestinal motility, visceral hypersensitivity, and the hypothalamic-pituitary-adrenal (HPA) axis are pivotal in this relationship (23,24). Stress, known to exacerbate IBS symptoms, is mediated through the HPA axis, indicating a direct link between psychological states and gastrointestinal function (23–25).

Moreover, psychiatric disorders and IBS demonstrate bidirectional comorbidities, with a significant overlap in symptomatology influenced by psychosocial stressors and affective factors (25,26). This bidirectionality is further supported by brain imaging studies showing morphological differences in IBS patients, particularly in areas related to pain modulation and emotional processing (27). For instance, changes in gray matter density in regions involved in cognitive and pain processing functions are observed in IBS patients, aligning with the heightened sensitivity to pain and stress (28,29).

These findings suggest that the mechanisms underlying the association between IBS and psychiatric disorders may involve alterations in brain structures and functions that modulate pain and emotional responses (23–28). Understanding these mechanisms can provide valuable insights into the causal pathways suggested by our study, helping to develop more effective treatment strategies that address both gastrointestinal and psychiatric aspects of IBS.

Our results indicating a reduced risk of IBS in patients with ADHD are intriguing and somewhat at odds with existing research. Specifically, Kedet et al.’s study reported a significant Odds Ratio (OR) of 1.67, suggesting an increased likelihood of IBS in ADHD patients (30). Similarly, Yeh et al.’s population-based study also found a significant association between ADHD and IBS (31). Our findings, however, point in the opposite direction, suggesting a potential decrease in IBS risk among those with ADHD. This divergence might be attributed to the different methodologies used (13). MR analysis, relying on genetic variants as instruments, offers insights into the genetic basis of these associations, which may not be captured fully in observational studies.

### Strengths and Limitations of the Study

One of the principal strengths of our study lies in the application of Two-Sample Mendelian Randomization (TSMR), which offers a robust approach to infer causal relationships by addressing common issues of confounding and reverse causation often encountered in observational studies (12). The use of extensive GWAS datasets and a variety of MR methods (IVW, MR Egger, weighted median) to ensure reliable results, further validated by sensitivity analyses and the MR regression test (13). However, limitations include the primary use of European ancestry data, potentially limiting wider applicability and the possibility of residual pleiotropy (32). Additionally, while we focused on genetic associations, this doesn’t encompass all factors, such as environmental or lifestyle influences, impacting these diseases.

### Conclusion and future research directions

Our study employs Mendelian Randomization to uncover a bidirectional causal association between IBS and MDD, and a unidirectional link with Anxiety Disorders. We found no significant connections with other psychiatric conditions such as Bipolar Disorder or Schizophrenia. These results support the substantial influence of the gut-brain axis in IBS and suggest that while IBS may predispose individuals to certain mental health issues, the converse relationship is not as clear. This underlines the importance of further research into the genetic and environmental underpinnings of IBS to inform holistic treatment strategies that address its complex ties to mental health.

## Data Availability

All data produced in the present study are available upon reasonable request to the authors

## Acknowledgment

none

## References

1. Saha L. Irritable bowel syndrome: Pathogenesis, diagnosis, treatment, and evidence-based medicine. World J Gastroenterol. 2014;20(22):6759.

2. Videlock EJ, Chang L. Latest Insights on the Pathogenesis of Irritable Bowel Syndrome. Gastroenterol Clin North Am. 2021 Sep;50(3):505–22.

3. Black CJ, Ford AC. Global burden of irritable bowel syndrome: trends, predictions and risk factors. Nat Rev Gastroenterol Hepatol. 2020 Aug 15;17(8):473–86.

4. Lee YT, Hu LY, Shen CC, Huang MW, Tsai SJ, Yang AC, et al. Risk of Psychiatric Disorders following Irritable Bowel Syndrome: A Nationwide Population-Based Cohort Study. PLoS One. 2015 Jul 29;10(7):e0133283.

5. Mohammed AA, Moustafa HA, Nour-Eldein H, Saudi RA. Association of anxiety-depressive disorders with irritable bowel syndrome among patients attending a rural family practice center: a comparative cross-sectional study. Gen Psychiatr. 2021 Dec 13;34(6):e100553.

6. Khan EH, Ahamed F, Karim MR, Roy P, Ahammed SU, Moniruzzaman M, et al. Psychiatric Morbidity in Irritable Bowel Syndrome. Mymensingh Med J. 2022 Apr;31(2):458–65.

7. Kawoos Y, Wani ZA, Kadla SA, Shah IA, Hussain A, Dar MM, et al. Psychiatric Co-morbidity in Patients With Irritable Bowel Syndrome at a Tertiary Care Center in Northern India. J Neurogastroenterol Motil. 2017 Oct 30;23(4):555–60.

8. Yeh HW, Chien WC, Chung CH, Hu JM, Tzeng NS. Risk of psychiatric disorders in irritable bowel syndrome-A nationwide, population-based, cohort study. Int J Clin Pract. 2018 Jul;72(7):e13212.

9. Schauer B, Grabe HJ, Ittermann T, Lerch MM, Weiss FU, Mönnikes H, et al. Irritable bowel syndrome, mental health, and quality of life: Data from a populationLJbased survey in Germany (SHIPLJTrendLJ0). Neurogastroenterology & Motility. 2019 Mar 15;31(3).

10. Popa SL, Dumitrascu DL. Anxiety and IBS RevisitedLJ: ten years later. Med Pharm Rep. 2015 Jul 22;88(3):253–7.

11. Talley NJ, Howell S, Poulton R. The Irritable Bowel Syndrome and Psychiatric Disorders in The Community: Is There A Link? American Journal of Gastroenterology. 2001 Apr;96(4):1072–9.

12. Minelli C, Del Greco M. F, van der Plaat Da, Bowden J, Sheehan NA, Thompson J. The use of two-sample methods for Mendelian randomization analyses on single large datasets. Int J Epidemiol. 2021 Nov 10;50(5):1651–9.

13. Sanderson E, Glymour MM, Holmes M V., Kang H, Morrison J, Munafò MR, et al. Mendelian randomization. Nature Reviews Methods Primers. 2022 Feb 10;2(1):6.

14. Liu B, Ye D, Yang H, Song J, Sun X, He Z, et al. Assessing the relationship between gut microbiota and irritable bowel syndrome: a two-sample Mendelian randomization analysis. BMC Gastroenterol. 2023 May 12;23(1):150.

15. Verbanck M, Chen CY, Neale B, Do R. Detection of widespread horizontal pleiotropy in causal relationships inferred from Mendelian randomization between complex traits and diseases. Nat Genet. 2018 May 23;50(5):693–8.

16. Nikolova VL, Pelton L, Moulton CD, Zorzato D, Cleare AJ, Young AH, et al. The Prevalence and Incidence of Irritable Bowel Syndrome and Inflammatory Bowel Disease in Depression and Bipolar Disorder: A Systematic Review and Meta-Analysis. Psychosom Med. 2022 Apr;84(3):313–24.

17. Fond G, Loundou A, Hamdani N, Boukouaci W, Dargel A, Oliveira J, et al. Anxiety and depression comorbidities in irritable bowel syndrome (IBS): a systematic review and meta-analysis. Eur Arch Psychiatry Clin Neurosci. 2014 Dec 6;264(8):651–60.

18. Hagberg KW, Li L, Peng M, Shah K, Paris M, Jick S. Incidence rates of suicidal behaviors and treated depression in patients with and without psoriatic arthritis using the Clinical Practice Research Datalink. Mod Rheumatol. 2016 Feb;26(5):774–9.

19. Garakani A, Win T, Virk S, Gupta S, Kaplan D, Masand PS. Comorbidity of Irritable Bowel Syndrome in Psychiatric Patients: A Review. Am J Ther. 2003 Jan;10(1):61–7.

20. Gupta S, Masand PS, Kaplan D, Bhandary A, Hendricks S. The relationship between schizophrenia and irritable bowel syndrome (IBS). Schizophr Res. 1997 Feb;23(3):265–8.

21. Mykletun A, Jacka F, Williams L, Pasco J, Henry M, Nicholson GC, et al. Prevalence of mood and anxiety disorder in self reported irritable bowel syndrome (IBS). An epidemiological population based study of women. BMC Gastroenterol. 2010 Dec 5;10(1):88.

22. Diao Z, Xu W, Guo D, Zhang J, Zhang R, Liu F, et al. Causal association between psycho-psychological factors, such as stress, anxiety, depression, and irritable bowel syndrome: Mendelian randomization. Medicine. 2023 Aug 25;102(34):e34802.

23. Mayer EA, Tillisch K. The Brain-Gut Axis in Abdominal Pain Syndromes. Annu Rev Med. 2011 Feb 18;62(1):381–96.

24. Posserud I. Altered visceral perceptual and neuroendocrine response in patients with irritable bowel syndrome during mental stress. Gut. 2004 Aug 1;53(8):1102–8.

25. Kim SE, Chang L. Overlap between functional GI disorders and other functional syndromes: what are the underlying mechanisms? Neurogastroenterology & Motility. 2012 Oct 2;24(10):895–913.

26. Surdea-Blaga T. Psychosocial determinants of irritable bowel syndrome. World J Gastroenterol. 2012;18(7):616.

27. Ellingson BM, Mayer E, Harris RJ, Ashe-McNally C, Naliboff BD, Labus JS, et al. Diffusion tensor imaging detects microstructural reorganization in the brain associated with chronic irritable bowel syndrome. Pain. 2013 Sep;154(9):1528–41.

28. Seminowicz DA, Labus JS, Bueller JA, Tillisch K, Naliboff BD, Bushnell MC, et al. Regional Gray Matter Density Changes in Brains of Patients With Irritable Bowel Syndrome. Gastroenterology. 2010 Jul;139(1):48–57.e2.

29. Blankstein U, Chen J, Diamant NE, Davis KD. Altered Brain Structure in Irritable Bowel Syndrome: Potential Contributions of Pre-Existing and Disease-Driven Factors. Gastroenterology. 2010 May;138(5):1783–9.

30. Kedem S, Yust-Katz S, Carter D, Levi Z, Kedem R, Dickstein A, et al. Attention deficit hyperactivity disorder and gastrointestinal morbidity in a large cohort of young adults. World J Gastroenterol. 2020 Nov 14;26(42):6626–37.

31. Yeh TC, Bai YM, Tsai SJ, Chen TJ, Liang CS, Chen MH. Risks of Major Mental Disorders and Irritable Bowel Syndrome among the Offspring of Parents with Irritable Bowel Syndrome: A Nationwide Study. Int J Environ Res Public Health. 2021 Apr 28;18(9):4679.

32. Mahajan A, Go MJ, Zhang W, Below JE, Gaulton KJ, Ferreira T, et al. Genome-wide trans-ancestry meta-analysis provides insight into the genetic architecture of type 2 diabetes susceptibility. Nat Genet. 2014 Mar 9;46(3):234–44.

